# Reconstructing the COVID-19 epidemic in Delhi, India: infection attack rate and reporting of deaths

**DOI:** 10.1101/2021.03.23.21254092

**Authors:** Margarita Pons-Salort, Jacob John, Oliver J Watson, Nicholas F Brazeau, Robert Verity, Gagandeep Kang, Nicholas C Grassly

**Affiliations:** MRC Centre for Global Infectious Disease Analysis; and the Abdul Latif Jameel Institute for Disease and Emergency Analytics (J-IDEA), School of Public Health, Imperial College London, UK; Department of Community Health, Christian Medical College, Vellore, India; Department of Gastrointestinal Sciences, Christian Medical College, Vellore, India

## Abstract

India reported over 10 million COVID-19 cases and 149,000 deaths in 2020. To estimate exposure and the potential for further spread, we used a SARS-CoV-2 transmission model fit to seroprevalence data from three serosurveys in Delhi and the time-series of reported deaths to reconstruct the epidemic. The cumulative proportion of the population estimated infected was 48.7% (95% CrI 22.1% – 76.8%) by end-September 2020. Using an age-adjusted overall infection fatality ratio (IFR) based on age-specific estimates from mostly high-income countries (HICs), we estimate that 15.0% (95% CrI 9.3% – 34.0%) of COVID-19 deaths were reported. This indicates either under-reporting of COVID-19 deaths and/or a lower age-specific IFR in India compared with HICs. Despite the high attack rate of SARS-CoV-2, a third wave occurred in late 2020, suggesting that herd immunity was not yet reached. Future dynamics will strongly depend on the duration of immunity and protection against new variants.

## Main text

With just under 150,000 COVID-19 deaths reported in 2020, India has a much lower reported COVID-19 mortality per million people than many other countries, such as Spain, France, the UK and the US. This discrepancy may be partly due to a younger population, but also incomplete documentation of deaths and of COVID-19 as a cause of death^1,2^. Assessing the extent of under-reporting of COVID-19 cases and deaths is essential to estimate the true burden of COVID-19 and likely future trends in transmission.

Multiple SARS-CoV-2 seroprevalence surveys conducted during 2020 in Delhi offer an opportunity to reconstruct the epidemic, assess the completeness of COVID-19 death reporting and estimate the infection attack rate in one of India’s largest metropolitan areas, home to 20 million people. SARS-CoV-2 transmission in Delhi has led to three waves of infection and mortality (Figure 1). At the beginning of the epidemic, all SARS-CoV-2 testing relied on RT-PCR, but since mid-June, antigen-based rapid diagnostic tests (Ag-RDTs), which have a lower sensitivity, have also been used and quickly exceeded the daily number of RT-PCR tests (Figure S1). Three serosurveys were conducted in Delhi in July, August and September, sampling individuals over 4 years old, found an age- and sex-adjusted seropositivity rate (uncorrected for test sensitivity and specificity) of 22.8%, 28.7% and 25.1% respectively (details in Table S1)^3^. Although the first serosurvey found a difference between slum and non-slum areas (25.3% vs. 19.2%, p<0.001), the second did not (28.9% vs. 28.8%, p=0.94), and the third did not report this information.

**Figure 1.**
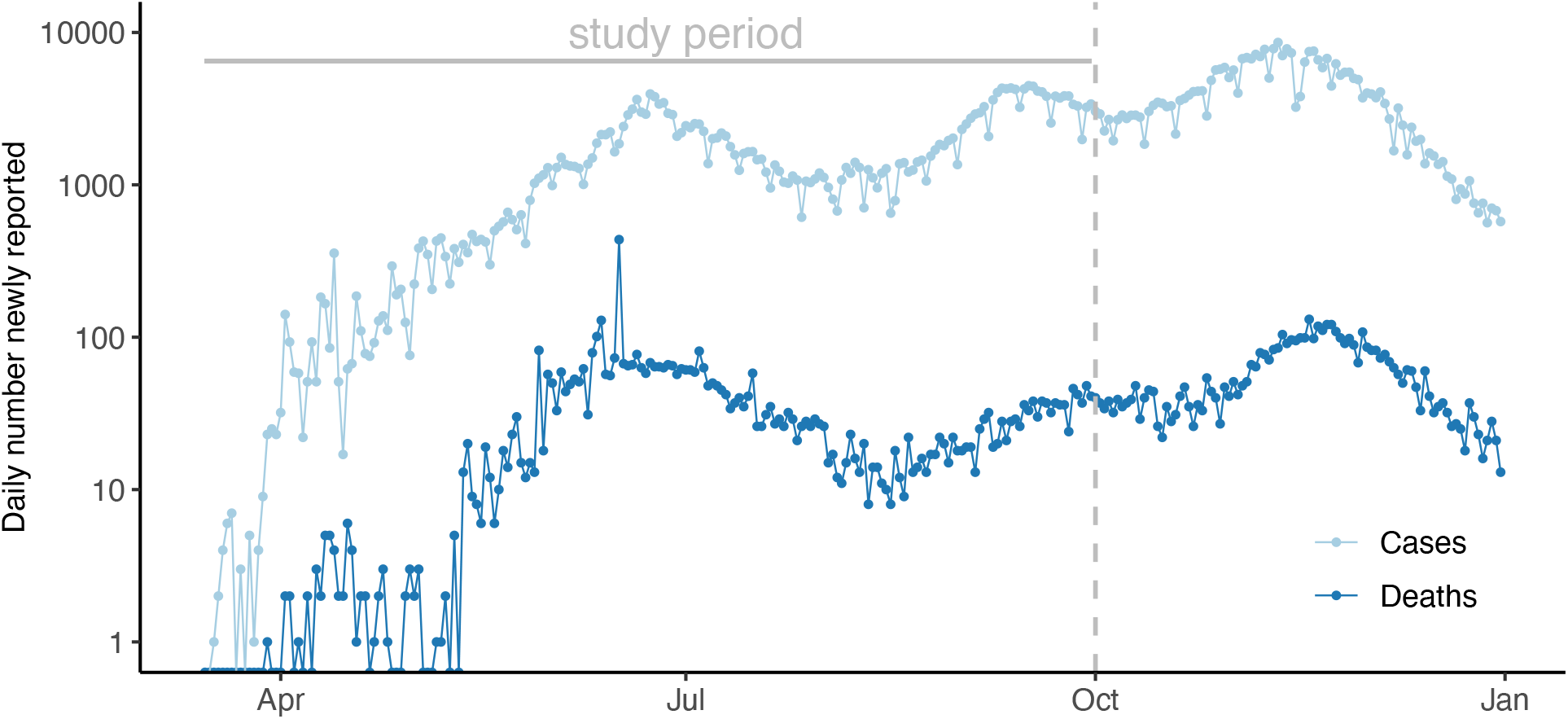
Epidemic of COVID-19 in Delhi. Daily number of COVID-19 reported cases (light blue) and deaths (dark blue) between the 15^th^ of March and the 31^st^ of December, 2020 in a logarithmic scale. The grey dashed vertical lines indicates the end of the study period (30^th^ of September 2020).

We developed a SARS-CoV-2 transmission model to estimate the incidence of infection and changes in the reproduction number (*R*) following implementation of non-pharmaceutical interventions, including lockdowns (Table S2, Figure S2). We used Bayesian Markov Chain Monte Carlo to fit the model to the three seroprevalence surveys and the time-series of reported deaths. The proportion of COVID-19 deaths reported was estimated by comparing reported deaths to those expected based on the age-adjusted IFR. We used estimates of the age-specific IFR based on data from 7 European countries, New York (USA) and Brazil^4^, to give a median age-adjusted IFR for Delhi of 0.39% (95% prediction interval 0.21 – 0.85%; this compares with ∼1% in high-income countries with older populations such as the UK^5,6^). Age-specific estimates from early data from China^6^ and from a meta-analysis in “advanced economies”^7^ gave a very similar value (0.39% and 0.40%, respectively). See the Online Methods for more details of the modelling and inference framework.

Our model fits the data well for both the time-series of deaths (Figure 2a) and seroprevalence survey data (Figure 2b), except the last serosurvey, where we estimate an increase in seropositivity with respect to the previous survey, instead of a slight decrease. This may be because the observation model does not account for waning antibodies and the possibility of seroreversion. However, the third serosurvey used a different testing kit, which could also contribute to this difference. We estimate that the first peak in incidence of infections was reached on the 31^st^ of May, when there were a median of 294,930 (95%CrI 143,271 – 440,702) new infections each day (Figure S4). The incidence at the second peak, reached on the 17^th^ of September, was slightly smaller, with a median of 79,032 (95%CrI 40,484 – 109,140) new infections per day. Assuming that transmission changes occur at the different changes in interventions and accounting for the depletion of susceptibles, we estimate that the effective reproduction number, *R*_*eff*_, increased during the relaxations introduced at phase 3 of the lockdown (starting on May 4), then decreased and then increased again in August (Figure 3a), resulting in a median infection attack rate of 48.7% (95% CrI 22.1% – 76.8%) by the end of September. Since then, Delhi has experienced a large third wave of cases and deaths (Figure 1), therefore suggesting that even with half of the population having been infected, the herd immunity threshold was not yet reached at that time. Interestingly, a serosurvey conducted in January 2021 found a sex- and age-adjusted seroprevalence of 56.1%, reflecting this third wave of transmission and probably indicating a steep increase in the cumulative number of infections.

**Figure 2.**
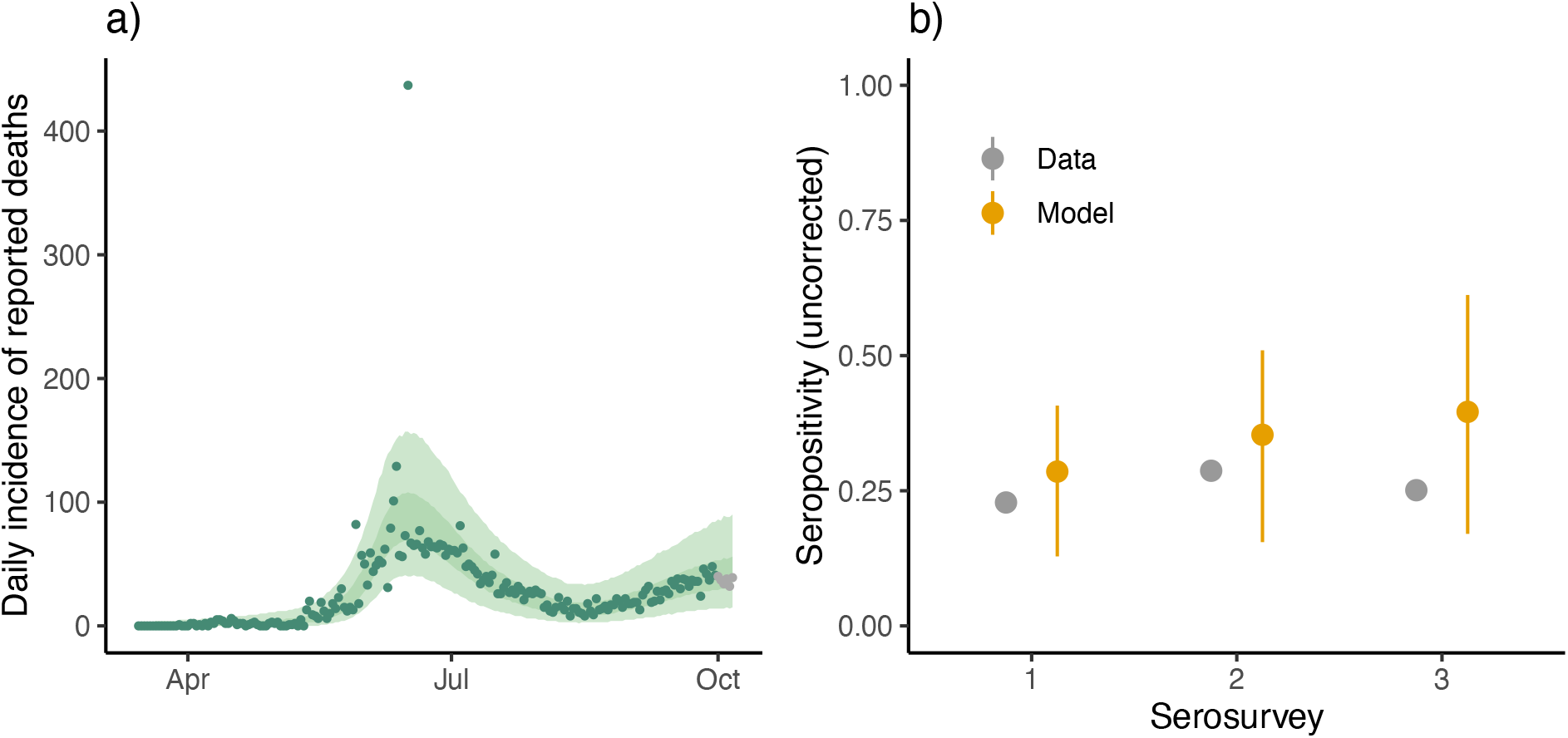
Model fit to data. (a) Model fit to the time-series of reported deaths (points), with 50% and 95% credible intervals (CrI). The last six points shown in gray were not used for parameter inference. (b) Median and 95% CrI model fit to seroprevalence data (gray) from three serosurveys.

**Figure 3.**
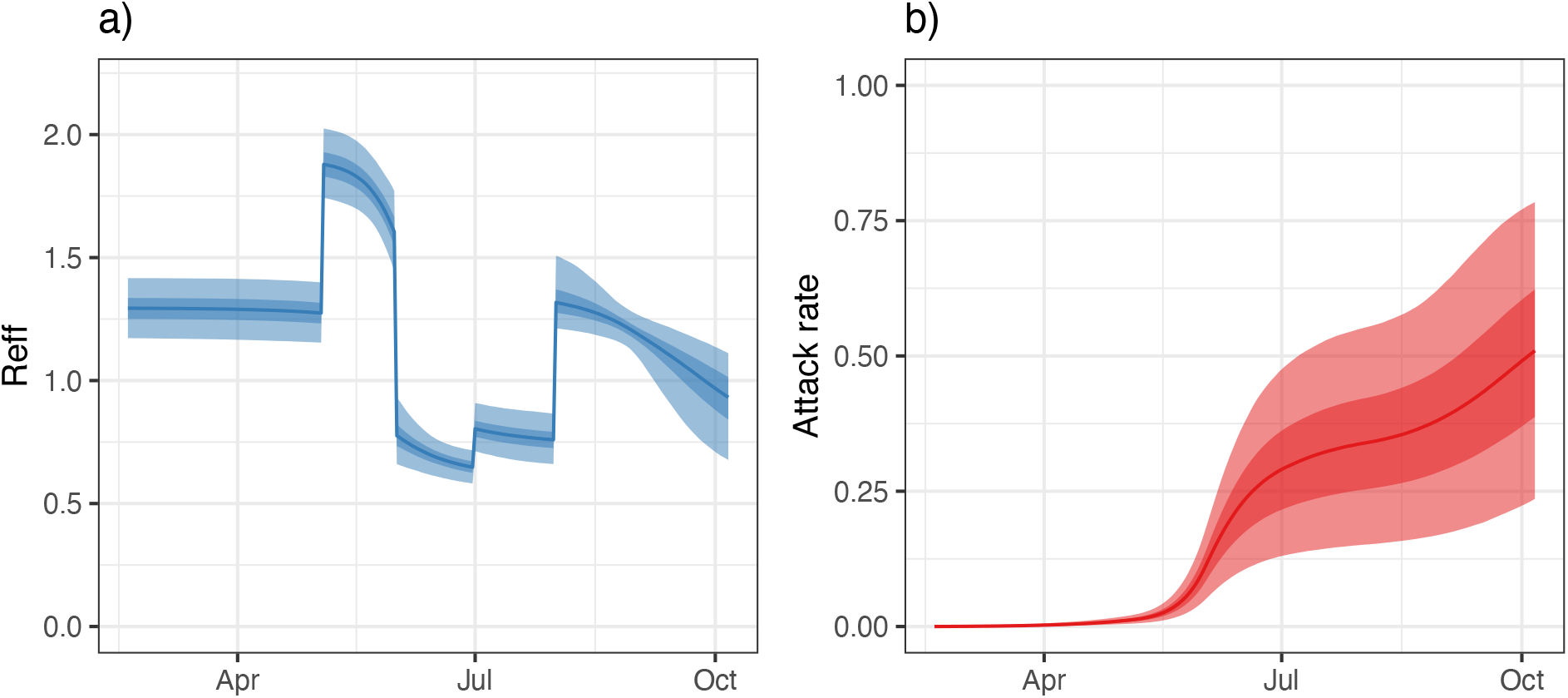
Effective reproduction number and attack rate. (a) Median and 50% and 95% CrI of the estimated reconstructed effective reproduction number. Changes are assumed to occur when changes in the interventions were introduced. (b) Median and 50% and 95% CrI of the estimated reconstructed infection attack rate.

Using the age-adjusted IFR of 0.39%, we estimate the reporting of deaths to be 15.0% (95% CrI 9.3% – 34.0%) (Figure S5). Repeating the analysis with an age-adjusted IFR of 0.21%, corresponding to the lower bound of the 95% prediction interval based on age-specific HIC data,^4^ increased the reporting rate to 28% (95% CrI 18 – 59%). This low rate of reporting is consistent with other cities in India where seroprevalence surveys suggest substantially greater exposure to infection than that predicted based on reported COVID-19 deaths. For example, comparison of seroprevalence during the first half of July 2020 in Mumbai^8^ with cumulative deaths at that time, gives an approximate estimate of reporting of 21% (Table S3). This high level of under-reporting may reflect both incomplete or delayed reporting of deaths and a failure to report COVID-19 as a suspected or confirmed cause of death, particularly in the absence of a SARS-CoV-2 test result. However, the extent of under-reporting is also dependent on the appropriateness of using an age-specific IFR in India derived from HIC data. The age-specific IFR may be lower in India for a number of reasons. First, the prevalence of comorbidities that increase the risk of severe COVID-19 following infection is somewhat lower in India than in the countries that informed the age-specific IFR estimates (Figure S6)^9^. However, correcting the Delhi IFR to account for the lower prevalence of comorbidities only marginally reduces the age-adjusted IFR (by up to 0.02%). Second, a recent study that analysed COVID-19 deaths by age from Mumbai and Karnataka found that the IFR rose less steeply with age than it did in high-income countries^10^. Third, differences in immunity reflecting exposure to a greater number of pathogens (including related coronaviruses) or simply lower frailty among those surviving to older ages in India compared with HICs could theoretically reduce the IFR in older groups, although data supporting these hypotheses are lacking.^11,12^.

Using the reconstructed incidence of infections, we also estimated the probability of detecting COVID-19 cases over time by comparing the number of reported cases to the estimated incidence of symptomatic infections (Figure 4a). The probability of infection detection quickly increased over the last weeks of March, fluctuated until mid-June, and remained relatively constant until the end of September, detecting a median of 7.1% of all symptomatic infections on average between July 1 and September 30, 2020 (Figure 4b).

**Figure 4.**
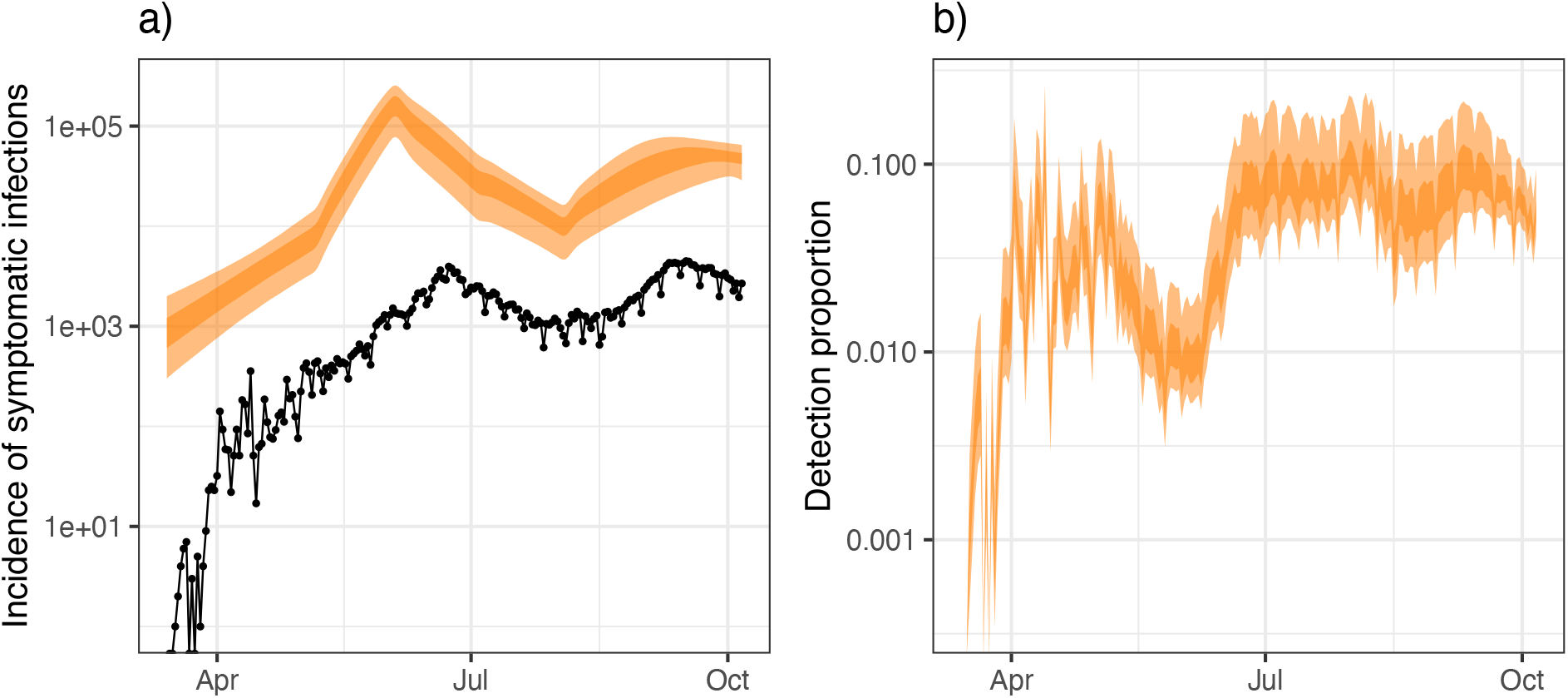
Detection of infections. (a) In orange, 50% and 95% CrI of the incidence of symptomatic infections, assuming that 2/3 of all infections are symptomatic. In black, the daily number of newly reported cases. (b) 50% and 95% CrI of the estimated detection probability per symptomatic infection per day.

This work has some limitations. First, the transmission model is not structured by age, and therefore, does not account for different mixing patterns between age classes and different attack rates by age. Nonetheless, age-structured models have predicted relatively flat infection attack rates across age for India^13^, consistent with age-stratified seroprevalence estimates^3^. This suggests that any bias in our results from age-specific patterns of mixing and potentially lower attack rates in more susceptible older age groups is likely to be limited. Second, we assume that reporting of deaths has been constant over the study period, but this value might have actually changed over time. Therefore our estimate of reporting represents an average over the study period. Finally, we use estimates of the age-specific IFR from mostly HICs and explore sensitivity to this assumption, including the use of data on comorbidities in India. Further analysis using data from cohort studies or demographic surveillance data in India will help to refine these estimates of the IFR and the exact degree of under-reporting of mortality.

At the time of writing (February 2021), the total number of new COVID-19 cases has been declining in India since mid-September 2020. How much of the country’s population has already been infected, and whether the herd immunity threshold has been reached are questions currently being debated^14^. Seroprevalence surveys conducted in major cities such as Mumbai, reported seroprevalence rates above 50% in slum areas for the first half of July^8^, suggesting that infection spread very quickly over the first few months in certain pockets, but below 20% in non-slums, showing the epidemic is highly heterogeneous in space. Therefore, the cumulative attack rate in rural areas and smaller cities may be far lower than that reached in major cities. Understanding what has brought the number of cases down in different places in India, and how to interpret the results from serosurveys in terms of the building up of population immunity is key to assess the future dynamics of the epidemic. Although a large proportion of the Indian population may have already been exposed, the circulation of new more transmissible or antigenically different variants, together with possible waning of population immunity over time, can result in the occurrence of new outbreaks.

## Methods

### Epidemiological and demographic data

Data on the number of SARS-CoV-2 confirmed cases and deaths reported daily in Delhi was available from the 14^th^ of March through the covid19india.org website. This is a volunteer-driven, crowdsourced initiative that collates data from several sources, including from the Ministry of Health and Family Welfare (MoHFW) and others. Cases and deaths that had occurred before the 14^th^ of March were reported as cumulative numbers on the first date of the dataset (i.e. 14^th^ March 2020). As we do not know when these cases and deaths occurred, we did not use the data reported on the 14^th^ of March for parameter inference.

We use data from three serosurveys conducted in Delhi^3^. The dates of sample collection, the number of samples tested, the seropositivity rate found, and the reported estimates of sensitivity and specificity of the assay used in each of the three serosurveys are summarised in Table S1.

We use projections of the population in Delhi for 2021 generated by the National Commission on Population^15^ split into age classes of 10 years.

### Transmission model

To model SARS-CoV-2 transmission we use a Susceptible-Exposed-Infected-Recovered (SEIR) deterministic transmission model (Figure S7, equations in the Supplementary Material). We do not stratify the population by age with respect to transmission parameters, and therefore assume random mixing by age such that the epidemic growth is equivalent in all age groups. We do not account for births, or deaths due to causes other than COVID-19, because of the short timeframe for the model. The generation time has been estimated at about 6.5 days, with infectiousness typically beginning in the day before symptoms start^16,17^. Given an incubation period of about 5.5 days,^18^ we therefore assumed a mean duration of the latency (pre-infectious period) of 4.5 days and a mean duration of infectiousness of 2 days to give the correct generation time.

### Disease progression and death model

We model disease progression and death following infection independently of the transmission process (Figure S7). As the model has been used for other purposes, it also includes hospitalisations, although these are not relevant for the work presented here and do not affect the results.

We use an incubation period (i.e. presymptomatic) with mean of 5.5 days, and a peaked distribution modelled by an Erlang distribution with shape parameter 6 as observed.^18^ We assume that one third of infections are asymptomatic (although there is high variability in the observed proportion across studies^19-21^ and a general increase in the proportion of infected that show symptoms through age^22,23^).

The proportion of total infections (asymptomatic and symptomatic) leading to hospitalisation and death are tracked separately – i.e. there is overlap between compartments for hospitalised individuals and those that will die (fatal infections). The proportion of infections leading to hospitalisation (with critical or non-critical care need respectively) are age-adjusted with the demographics from Delhi and age-stratified estimates from ^6^ based on data from China. Similarly, the proportion of infections leading to death (i.e. the age-adjusted IFR) is based on estimates of age-stratified IFR from^4^ applied to the population of Delhi.

The average time from symptom onset to hospitalisation is set to 5.8 days, consistent with observations in China^24^. We assume a mean duration of hospital stay of 9.8 days if no critical care and 9.8 days if critical care required, followed by 3.3 days to recover in non-critical care (stepdown), based on early, unpublished UK estimates. Note these estimates do not affect our results, which are not based on hospitalisations. The average time from symptom onset to death was around 16 days^6^. We therefore assume a mean delay between the time of hospitalisation and death of 10 days. These values may be different for India, but no data is currently available.

### Parameter inference

We fit the transmission model to both the seroprevalence data and the daily incidence of COVID-19 deaths reported between the 15^th^ of March and the 30^th^ of September, 2020. To account for under-reporting and overdispersion on the death data, we model the number of deaths with a Negative Binomial distribution:

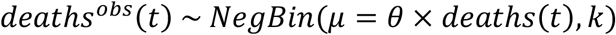

where the mean is *θ* × *deaths*(*t*) and the variance is *θ* × *deaths*(*t*) + (*θ* × *deaths*(*t*))^2^/(*θ* × *deaths*(*t*))^*k*^.

We model the number of individuals that would test seropositive each day with a given serological assay of sensitivity *Se*_*j*_ and specificity *Sp*_*j*_ as

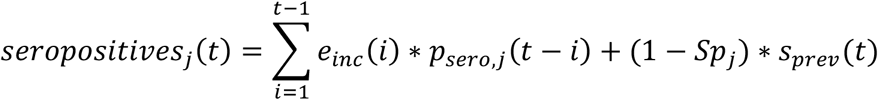

where *e*_*inc*_ is the incidence of infection, *p*_*sero,j*_(τ) is the probability of testing positive τ days after infection and *s*_*prev*_ is the number of susceptible, as in Ojal et al.^25^. We assume that *p*_*sero,j*_(τ) increases linearly from 0 the day of infection to *Se*_*j*_ 26 days after infection and remains constant after that (i.e. we do not consider seroreversion):

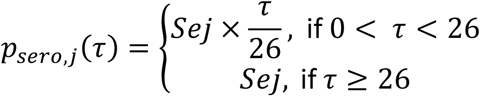

Finally, we link the modelled number of seropositives at the mid time point of each serosurvey (denoted here by ts_1_, ts_2_ and ts_3_ respectively for the serosurveys 1, 2 and 3) to the data from the three seroprevalence surveys using Beta Binomial distributions to account for overdispersion:

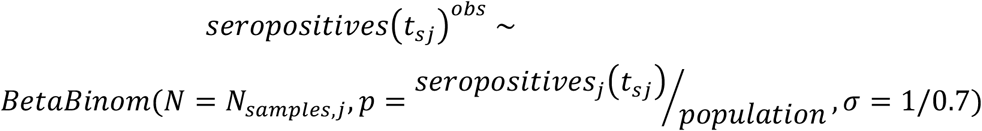

with the overdispersion parameter fixed to σ = 1/0.7, due to the small number of observations available to accurately estimate it.

We allow the reproduction number, *R*, to change at 5 different time points corresponding to the time of changes in interventions (Table S2). We denote *R*_0_ the basic reproduction number during the first period (i.e. before any changes), and *R*_*i*_ the reproduction number after the i-th change and during the (i+1)-th (i in 1,…,5) period. We parameterise it as *R*_*i*_ = *R*_0_ × (1 + *r*_1_) × … × (1 + *r*_*i*_).

We estimate the initial value of the reproduction number (*R*_0_) and the subsequent changes at each time point (*r*_1_, …, *r*_5_), the initial number of infected (*E*(0) + *I*(0)), the reporting (*θ*) and overdispersion of deaths (*K*). We assume the starting time for the simulations, *t*_0_, to be the 19^th^ of February 2020 (i.e. 28 days before the first ten cases were reported) and estimate the number of infected individuals at that time point (*E*(0) + *I*(0)).

The first change on the reproduction number, corresponding to the start of the lockdown on the 25^th^ of March (and modelled through the parameter *r*_1_), could not be estimated, because the number of deaths at that time did not allow to infer a change in transmission (no deaths reported between the 15^th^ and the 28^th^ of March). We therefore assumed *r*_1_ = 0, and the subsequent change on the reproduction number (*r*_2_) was assumed to occur on the 4^th^ of May, when the first relaxations were introduced. Therefore, the estimate of the reproduction number between the beginning of the simulations (19^th^ of February 2020) and until the first estimated change on the 4^th^ of May 2020 implicitly accounts for any effects of the lockdown at that time.

Because *R*_0_ and the initial number of infected are highly correlated, we estimate the total number of infections just before the first change on the reproduction number, on the 4^th^ of May, and back-calculate the initial number of infected using the relationship given by a simple exponential growth model and the relationship between *R*_0_ and the epidemic growth rate for an SEIR model ^26^, as in Salje et al. ^27^.

Model parameters were estimated using Markov Chain Monte Carlo via the lazymcmc package^28^ with 100,000 iterations and uniform prior distributions. Four chains with different starting values were run to check convergence.

All the analyses were implemented in R 4.0.2 ^29^.

## Data Availability

All data used in the manuscript is publicly available.

https://www.covid19india.org

## Acknowledgements

We thank Nimalan Arinaminpathy for insightful comments on the manuscript, Marc Baguelin for helpful discussions on parameter inference and James A. Hay for help using the lazymcmc R package. M.P.-S. is a Sir Henry Dale Fellow jointly funded by the Wellcome Trust and the Royal Society (grant number 216427/Z/19/Z). M.P.-S., O.J.W., N.F.B., R.V., and N.C.G. acknowledge funding from the MRC Centre for Global Infectious Disease Analysis (MR/R015600/1), jointly funded by the UK Medical Research Council (MRC) and the UK Foreign, Commonwealth & Development Office (FCDO), under the MRC/FCDO Concordat agreement and is also part of the EDCTP2 programme supported by the European Union; and acknowledge funding by Community Jameel.

## Author contributions

M.P.-S. and N.C.G. designed the study. M.P.-S. performed the analyses and wrote the first draft of the manuscript. N.B. and R.V. provided code for the analysis. All authors critically reviewed the methods, and contributed to the interpretation of the results and writing of the manuscript.

## Competing interests

The authors declare no competing interests.

## Notes

### Competing Interest Statement

The authors have declared no competing interest.

### Author Declarations

No necessary IRB.

