## Supplementary Information for "Reconstructing the COVID-19 epidemic in Delhi, India: infection attack rate and reporting of deaths"

**Equations for the transmission model.** See notation and description of variables and parameters in Tables S4 and S5 respectively.

$$\frac{dS}{dt} = -\lambda S$$

$$\frac{dE}{dt} = \lambda S - \omega E$$

$$\frac{dI}{dt} = \omega E - \gamma I$$

$$\frac{dR}{dt} = \gamma I$$

$$\lambda = R_i \gamma \frac{I}{N}$$

$$N = S + E + I + R$$

**Equations for the disease-progression model.** See notation and description of variables and parameters in Tables S4 and S5 respectively.

$$\frac{dPSY_1}{dt} = \lambda S - \frac{6}{D_{PSY}} PSY_1$$

$$\frac{dPSY_2}{dt} = \frac{6}{D_{PSY}} PSY_1 - \frac{6}{D_{PSY}} PSY_2$$

…

$$\frac{dPSY_6}{dt} = \frac{6}{D_{PSY}} PSY_5 - \frac{6}{D_{PSY}} PSY_6$$

$$\frac{d(SY + ASY)}{dt} = \frac{6}{D_{PSY}} PSY_6 - \frac{1}{D_{SY}} (SY + ASY)$$

$$\frac{dHNC}{dt} = p_{inf,hnc} \frac{1}{D_{SY}} (SY + ASY) - \frac{1}{D_{HNC}} HNC$$

$$\frac{dHC}{dt} = p_{inf,hc} \frac{1}{D_{SY}} (SY + ASY) - \frac{1}{D_{HC}} HC$$

$$\frac{dHSD}{dt} = p_{hc,hsd} \frac{1}{D_{HC}} HC - \frac{1}{D_{HSD}} HSD$$

$$\frac{dF}{dt} = p_{inf,death} \frac{1}{D_{SY}} (SY + ASY) - \frac{1}{D_F} F$$

$$\frac{dD}{dt} = \frac{1}{D_F} F$$

**Table S1.** Information on the three serosurveys conducted in Delhi[1] during the study period. Serosurveys 1 and 2 used the testing kit ELISA COVID-Kawach kit, whereas serosurvey 3 used the ERBALISA COVID-19 IgG.

| Serosurvey and body in charge | Date (mid time point) | Number of samples | Uncorrected seropositivity rate | Test Sensitivity | Test Specificity |
|---|---|---|---|---|---|
| 1, NCDC | 01/07/2020 | 19,041 | 0.2283 | 0.921 | 0.977 |
| 2, MAMC | 04/08/2020 | 15,046 | 0.287 | 0.921 | 0.977 |
| 3, MAMC | 04/09/2020 | 17,409 | 0.251 | 0.9912 | 0.9933 |

Abbreviations: NCDC, National Center for Disease Control; MAMC: Maulana Azad Medical College.

**Table S2.** National interventions to control COVID-19 in India.

| Intervention | Start date | End date | Modeled change in R? | Observations |
|---|---|---|---|---|
| Janata curfew | 22/03/2020 | | No | |
| Phase 1 lockdown | 25/03/2020 | 14/04/2020 | No[a] | Beginning of lockdown |
| Phase 2 lockdown | 15/04/2020 | 03/05/2020 | No | Lockdown extension |
| Phase 3 lockdown | 04/05/2020 | 17/05/2020 | Yes | Lockdown extension with relaxations |
| Phase 4 lockdown | 18/05/2020 | 31/05/2020 | No | Lockdown extension |
| Unlock 1.0 | 01/06/2020 | 30/06/2020 | Yes | Re-opening phase |
| Unlock 2.0 | 01/07/2020 | 31/07/2020 | Yes | Re-opening phase |
| Unlock 3.0 | 01/08/2020 | 31/08/2020 | Yes | Re-opening phase |
| Unlock 4.0 | 01/09/2020 | 30/09/2020 | No | Re-opening phase |

[a] Not enough data available to estimate a change in transmission at that point.

**Table S3.** Estimate of death reporting in Delhi and Mumbai based on a simple back calculation using data on the cumulative number of deaths up until the first serosurvey. For Mumbai, we use the seroprevalence data reported in [2], we assume that 53% of the population lives in slums, and account for different age distribution in slums and non-slums, based on data from the TIFR Covid-19 City-Scale Simulation Team[3].

| Area | Date[a] | Cumulative reported deaths | Seroprev.[b] | Population size | IFR[c] | Death reporting |
|---|---|---|---|---|---|---|
| **Delhi** | 01/07/2020 | 2,803 | 22.86% | 20.86M | 0.39% | 15% |
| **Mumbai** | 07/07/2020 | 3,317 | - | 12.8M | - | 21% |
| ***Slums*** | - | - | 55.7% | 6.784M | 0.29% | - |
| ***Non-slums*** | - | - | 16.2% | 6.016M | 0.51% | - |

[a] Mid time point of sample collection during the serosurvey.

[b] Corrected for test sensitivity and specificity.

[c] Based on the model in [4].

**Table S4.** Model variables.

| Notation | Description |
|---|---|
| **S** | Susceptible |
| **E** | Exposed |
| **I** | Infected |
| **R** | Recovered |
| **N** | Total population size |
| **PSY1,…,PSY6** | Pre-symptomatic 1,…, 6 (the 6 compartments are necessary to model the Erlang distribution with shape value 6) |
| **SY** | Symptomatic |
| **ASY** | Asymptomatic |
| **HNC** | Hospitalised non-critical care |
| **HC** | Hospitalised critical care |
| **HSD** | Hospitalised step-down |
| **F** | Fatal infections |
| **D** | Deaths |

**Table S5.** Model parameters.

| Notation | Description | Value and units |
|---|---|---|
| $R_0$ | Basic reproduction number | Estimated |
| **r1,…,r5** | Coefficients modifying the reproduction number | Estimated |
| $1/\omega$ | Mean duration latent period | 4.5 days |
| $1/\gamma$ | Mean duration infectious period | 2 days |
| $D_{PSY}$ | Mean duration pre-symptomatic | 5.5 days |
| $D_{SY}$ | Mean duration symptomatic | 5.8 days |
| $D_{HNC}$ | Mean duration hospitalised non critical care | 9.8 days |
| $D_{HC}$ | Mean duration hospitalised critical care | 9.8 days |
| $D_{HSD}$ | Mean duration hospitalised step-down | 3.3 days |
| $D_F$ | Mean duration infections leading to fatal outcome | 10 days |
| $p_{INF,HNC}$ | Proportion of infections leading to hospitalisation in non critical care (age-adjusted) | $2.77*10^{-2}$ |
| $p_{INF,HC}$ | Proportion of infections leading to hospitalisation in critical care (age-adjusted) | $6.64*10^{-3}$ |
| $p_{INF,D}$ | Proportion of infections leading to death (age-adjusted IFR) | $3.94*10^{-3}$ † |
| $p_{HC,HSD}$ | Proportion of hospitalisations in critical care recovering (i.e. going to step-down compartment) | 0.6 |

† Value used in the main analysis. Sensitivity analysis performed on this value.

**Figure S1.** Number of tests. Daily number of RT-PCR and Ag-RTD tests performed between the 14th of June and 16th of September, 2020. Ag-RTDs were not used before this period.

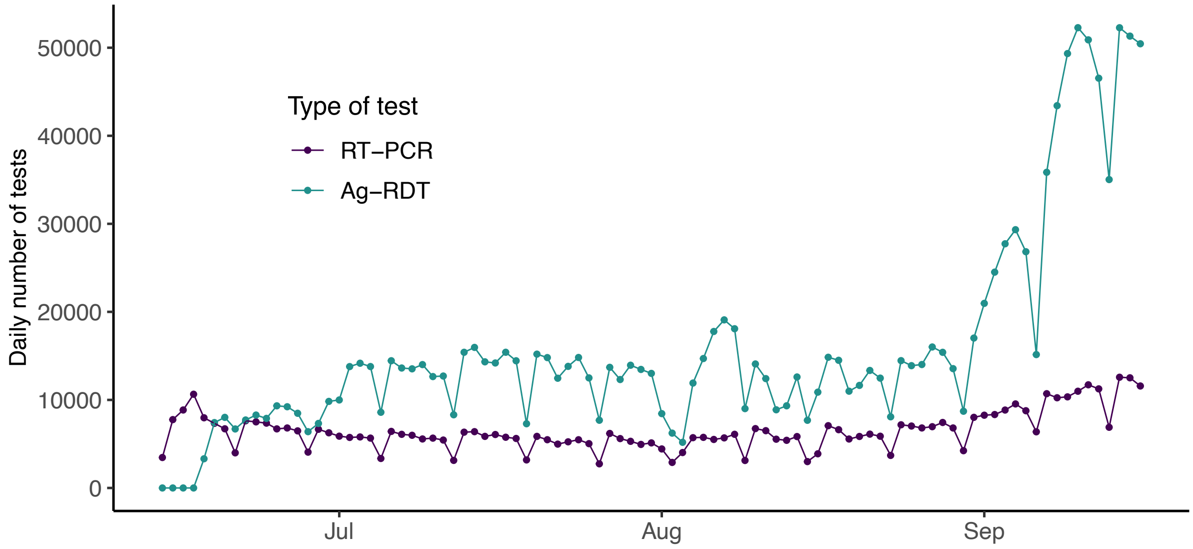

**Figure S2.** Changes in mobility over time based on summarised Google data between the 15th of February and 31st of December, 2020. The data have been smoothed to remove the weekend effect and averaged across five of the six data streams available (all, except residential, see Figure S3). Coloured lines indicate the start of interventions listed in Table S2. The grey vertical line indicates the end of the study period (30th of September 2020).

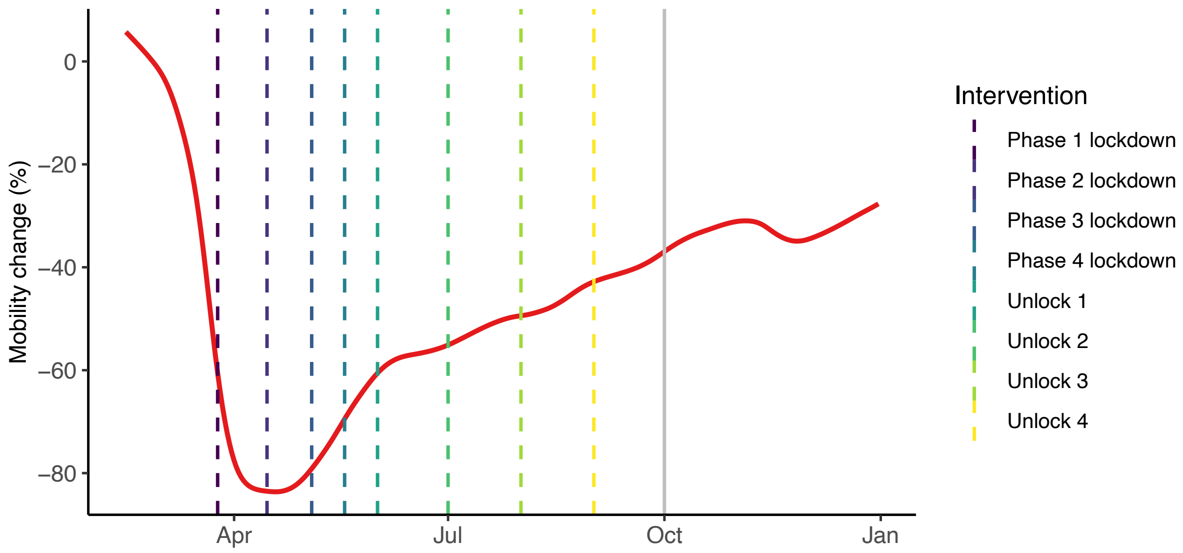

**Figure S3.** Google mobility data for Delhi. The raw data for the six data streams available from Google mobility are shown in red for retail and recreation, in blue for grocery and pharmacy, in green for parks, in purple for transit stations, in orange for workplaces and in yellow for residential. The dashed black lines are smoothed lines of the raw data obtained with a spline after removing the points for Thursday to Sunday to remove the weekend effect. These smoothed lines for all the streams, except for the residential one, were averaged to produce Figure S2. The vertical lines indicate the beginning of the lockdown, on the 25th of March 2020.

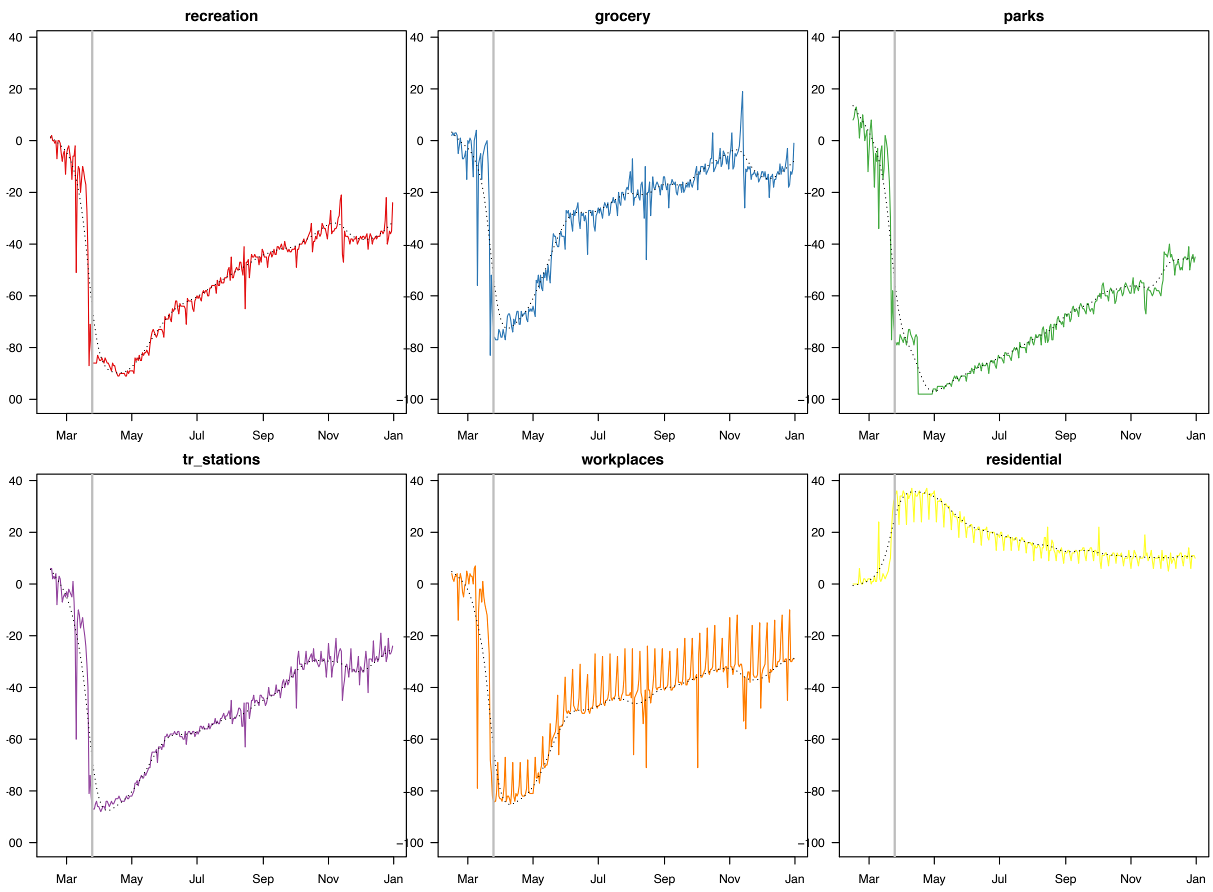

**Figure S4.** Reconstructed estimated incidence of infections.

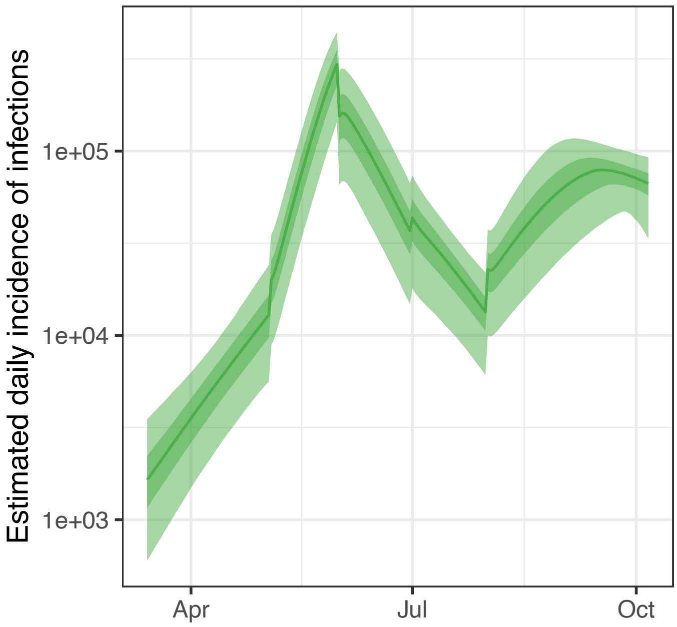

**Figure S5.** Posterior densities of estimated parameters for four different MCMC chains initialised at different starting values.

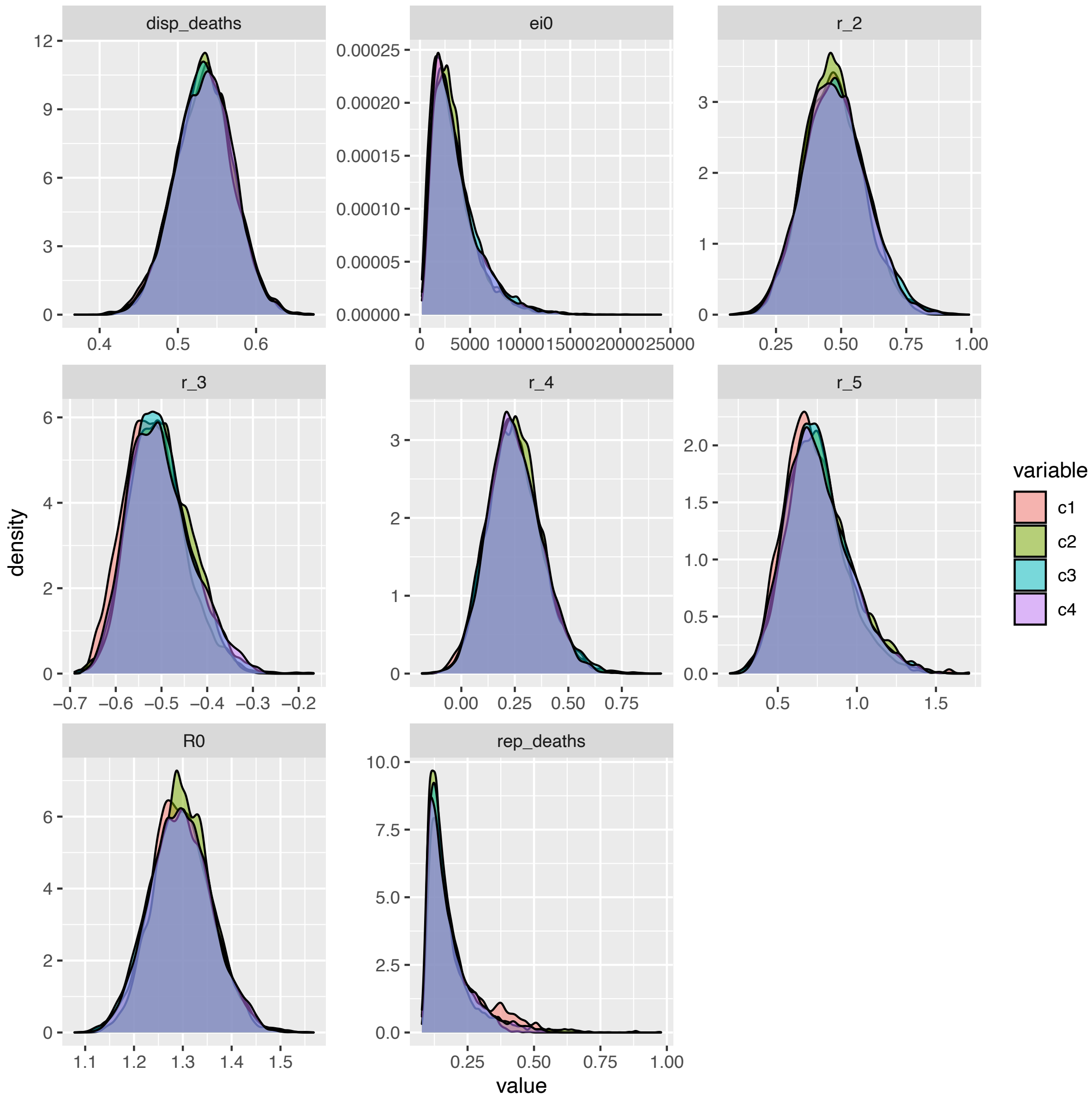

**Figure S6.** Prevalence of at least one or at least two co-morbidities for severe COVID-19 by age for India (black dotted line) and the nine countries that informed the age-specific estimates of the IFR (extracted from Clark et al[5]).

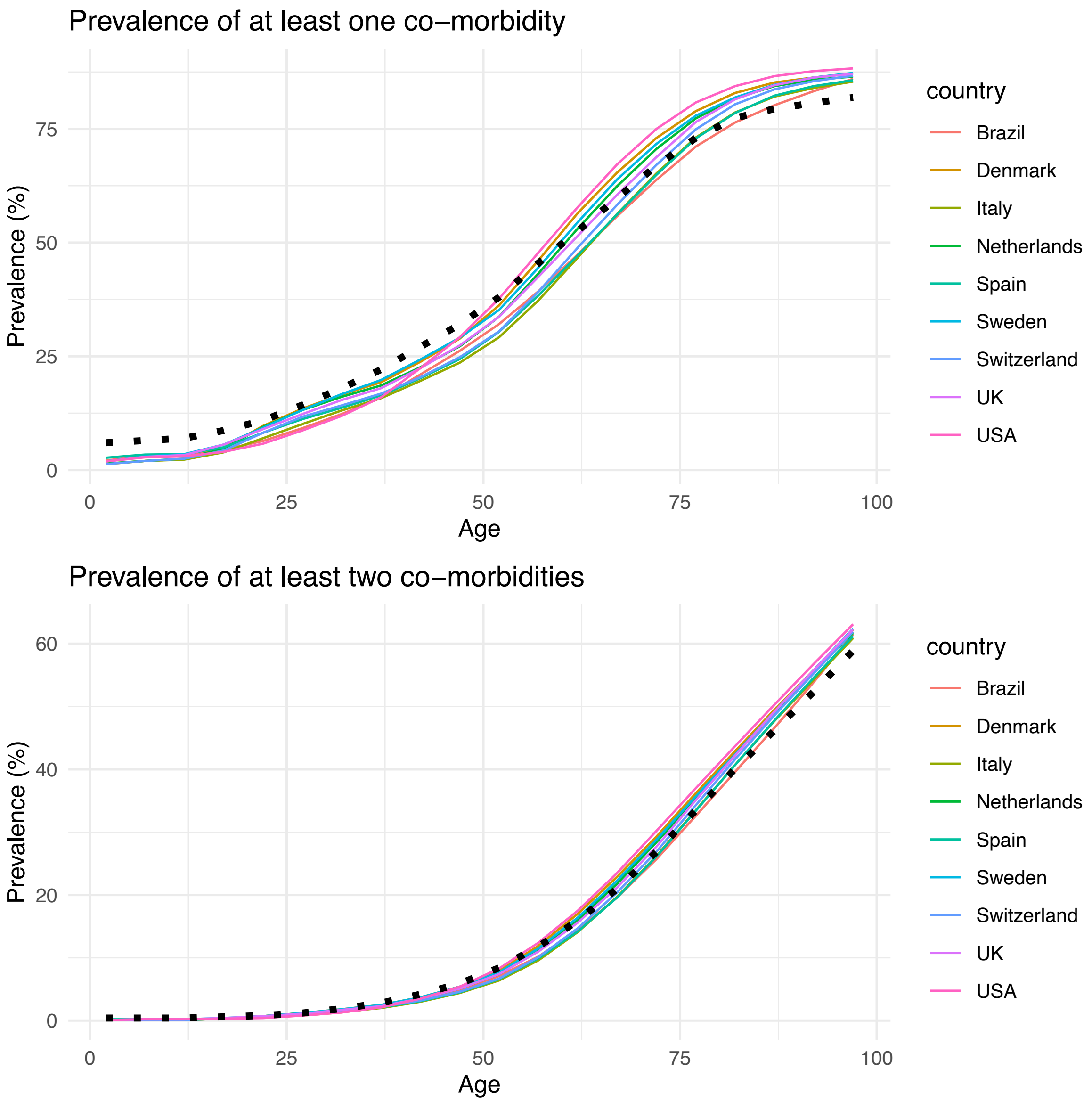

**Figure S7.** Model diagram. In green, the SEIR transmission model. In blue, the disease-progression model, with infections leading to death in red.

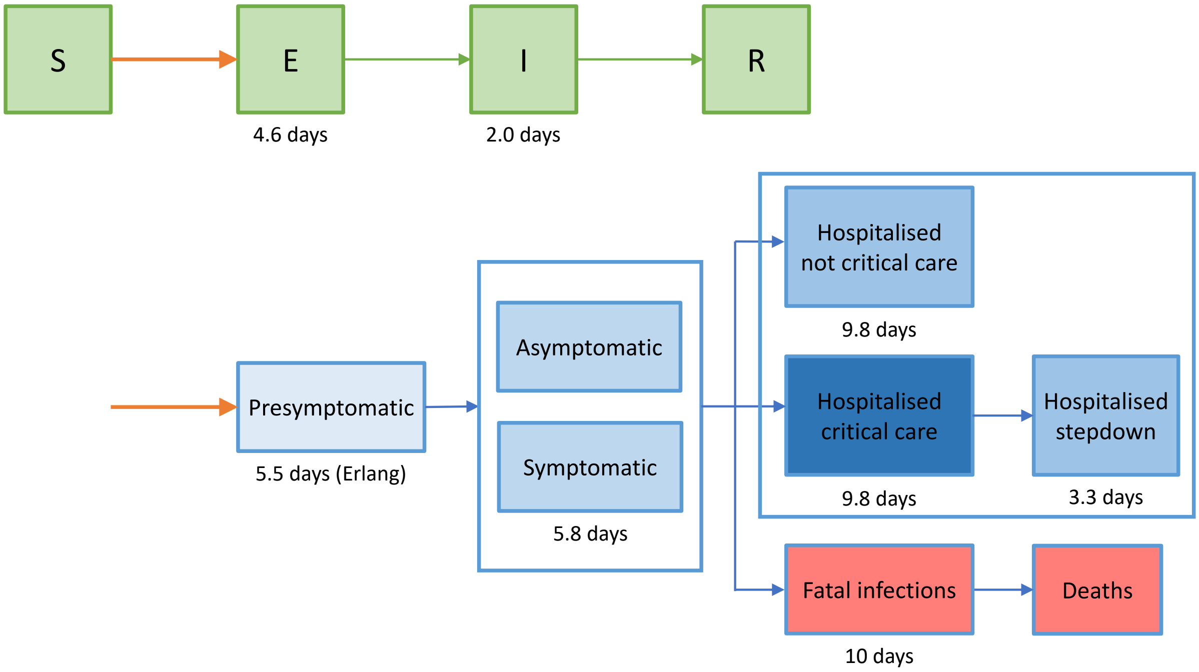
